# Efficacy and safety of antidepressants for the treatment of low back pain and osteoarthritis: protocol for a systematic review and meta-analysis

**DOI:** 10.1101/2020.04.30.20086827

**Authors:** Giovanni E Ferreira, Chung-Wei Christine Lin, Joshua Zadro, Christina Abdel-Shaheed, Andrew J McLachlan, Mary O’Keeffe, Chris G Maher

**Author notes:** Corresponding author, Giovanni E. Ferreira, Institute for Musculoskeletal Health, School of Public Health, Faculty of Medicine and Health, The University of Sydney, Sydney, New South Wales, Australia, PO Box M179, Missenden Road, Camperdown | NSW | 2050, Australia.

## Abstract

This preprint describes the protocol for a systematic review and meta-analysis evaluating the efficacy and safety of antidepressants for the treatment of low back pain and osteoarthritis. The review protocol was originally submitted to the International Prospective Register of Systematic Reviews (PROSPERO) on November 14 2019. At the time of the protocol submission, the review had not started yet. Electronic database searches were formally conducted on 15 November 2019, after we received an acknowledgement of receipt email (Appendix 3).

In correspondence with PROSPERO on April 9 2020, we were advised that PROSPERO staff were dealing with an unprecedented demand of systematic review protocols, and that COVID-19-related protocols were being given priority (Appendix 4). As a result, review protocol submissions outside the UK were expected to take longer than five months to be processed.

Following PROSPERO’s suggestion, we decided to deposit the review protocol on Open Science Framework as a preprint. This document contains the systematic review protocol originally registered on PROSPERO (Appendix 2) and evidence that confirms that the protocol was prospectively registered.

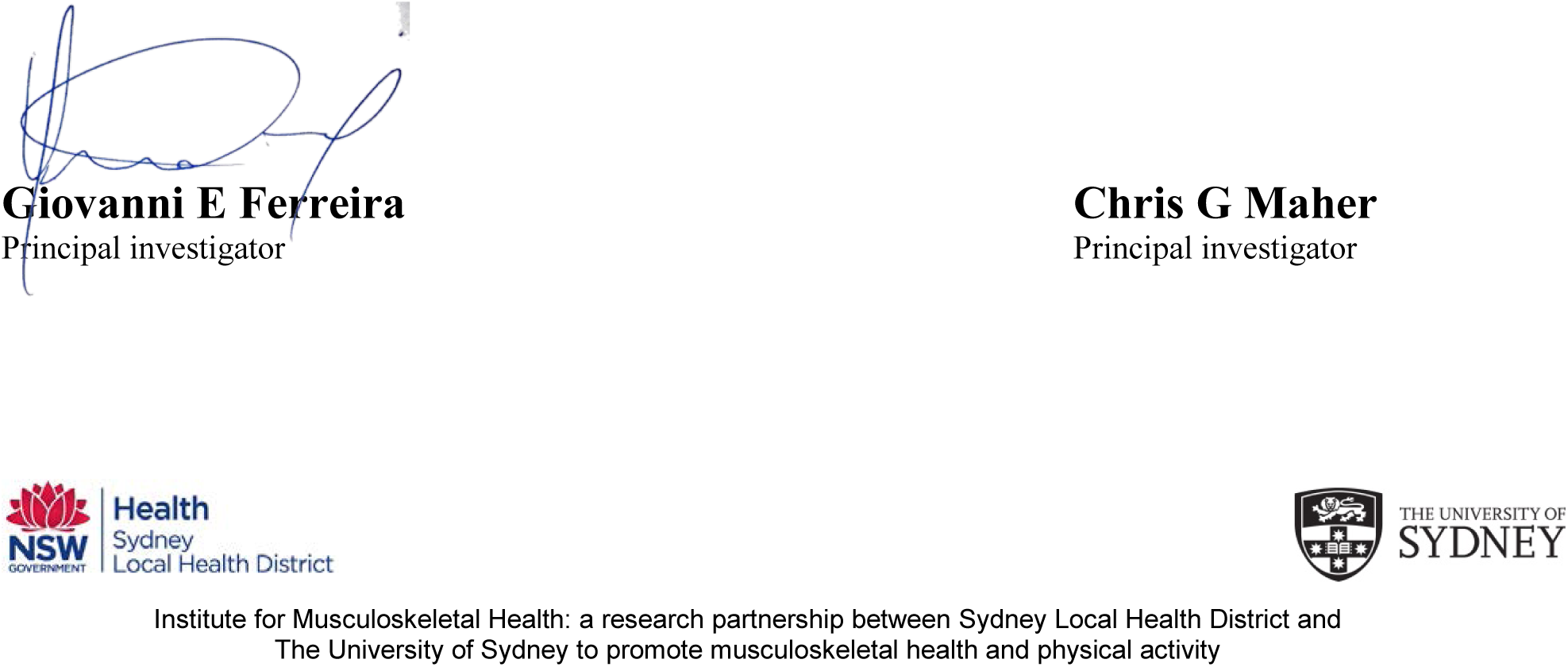

## EFFICACY AND SAFETY OF ANTIDEPRESSANTS FOR THE TREAMENT OF LOW BACK PAIN AND OSTEOARTHRITIS

### Review question

Are antidepressants efficacious and safe for patients with spinal pain or osteoarthritis?

### Searches

We will search MEDLINE (via Ovid), EMBASE (via Ovid), Cochrane Central Register of Controlled Trials, CINAHL, International Pharmaceutical Abstracts, ClinicalTrials.gov and the World Health Organization International Clinical Trials Registry Platform (ICTRP) from their inception to November 2019.

## Types of study to be included

Randomised placebo-controlled trials published as full-text in peer-reviewed journals. Both enriched and non-enriched designs will be included. We will contact authors who have published a protocol for a relevant trial that has not yet been published in an attempt to access the data or to obtain a copy of the publication. There will be no restrictions for language or publication date. We will attempt to translate studies published in languages other than English.

## Condition or domain being studied

Spinal pain (low back pain and/or neck pain), and hip and/or knee osteoarthritis (OA).

## Participants/population

Participants with a diagnosis of non-specific spinal pain (low back and/or neck pain), spinal pain with radicular symptoms and/or radiculopathy; and hip and/or knee OA. Studies with mixed populations will be included. Studies that include participants with serious spinal pathologies (e.g. cauda equina syndrome, cervical myelopathy, spinal tumours, spinal infection) will be excluded. We will exclude studies including participants with inflammatory arthritis eg axial spondyloarthritis, rheumatoid arthritis unless data for knee and/or hip OA is reported separately. Studies where participants received previous spinal or OA surgery are eligible, but studies evaluating immediate post-operative pain management (ie within past month) will not be included.

## Intervention(s), exposure(s)

Antidepressants are recommended by clinical practice guidelines for low back pain^1^ and hip and knee osteoarthritis.^2,3^ We will include trials testing any type of antidepressant drug prescribed at any dose, as treatment for spinal pain and/or hip and/or knee OA. Studies testing drug combinations (e.g. antidepressants in addition to other analgesics) will only be included if the treatment contrast is other analgesic in addition to placebo Types of antidepressants drugs to be included in this review include, but are not limited to:

- Tricyclic antidepressants (e.g. amitriptyline, nortriptyline, desipramine, imipramine, doxepin, dothiepin, trimipramine, clomipramine and protriptyline)
- Tetracyclic antidepressants (e.g. mianserin amoxapine and maprotiline)
- Selective serotonin reuptake inhibitors (SSRIs) (e.g. citalopram, escitalopram, fluoxetine, fluvoxamine, paroxetine, Zimelidine and sertraline)
- Serotonin and norepinephrine reuptake inhibitors (SNRIs) (e.g. Duloxetine, Venlafaxine, Desvenlafaxine, Milnacipran)
- Monoamine oxidase inhibitors (MAOIs) (e.g. harmaline, iproclozide, iproniazid, isocarboxazid, moclobemide nialamide, toloxatone and tranylcypromine)
- Atypical antidepressants (e.g. Bupropion, Trazodone, Mirtazapine, Nefazodone, agomelatine)

## Data Availability

Data for the full systematic review will publicly available.

## Comparator(s)/control

We will include both inert (e.g. an inert substance that does not contain an active drug treatment) and active placebos (i.e. drugs that have no known effect on pain but may mimic the adverse effects of antidepressants).

**Primary outcomes**

1. Pain intensity (e.g. numerical pain rating scale [NPRS], visual analogue scale [VAS]);
2. Disability (e.g. Oswestry Disability Index, Roland Morris Disability Questionnaire)
3. Composite disease scores (e.g Western Ontario and McMaster Universities osteoarthritis index [WOMAC], Knee Osteoarthritis Outcome Score [KOOS], Knee Society Scoring System [KSSS], Lysholm Knee Scoring Scale [LKSC], Hip Osteoarthritis Outcome Score [HOOS], Harris Hip Score [HHS])

Secondary outcomes
1. Adverse events

- Number of participants reporting any adverse events (as defined by each study)
- Number of participants reporting any serious adverse events (as defined by each study)
2. Number (%) of participants who stopped the study medicine due to adverse effects
3. Number (%) of participants who stopped the study medicine due to lack of efficacy
4. Number (%) of participants who withdrew from study citing either of above
5. Number (%) of participants who were lost to follow-up

**Timing and effect measures**

1. Immediate term (≤2 weeks);
2. Short term (>2 weeks but ≤ 3 months);
3. Intermediate term (>3 months but ≤12 months);
4. Long term (>12 months).

In studies with multiple time points, we will extract data from the time point closest to 2 weeks, 3, 6 or 12 months.

## Data extraction (selection and coding)

Two independent researchers will extract the following data using a standardised data extraction form: bibliometric data, study characteristics, characteristics of included participants, and outcome measures.

## Risk of bias (quality) assessment

We will assess risk of bias using the Cochrane Risk of Bias Tool. We will rate the quality of the evidence for each outcome measure using the GRADE framework. The domains evaluated by the GRADE framework include study design, risk of bias, inconsistency, indirectness, imprecision and publication bias. For each domain, the quality of evidence was downgraded by one level if a serious flaw was present using the following criteria:

- **Risk of bias**: downgrade by one level if more than 25% of participants are from studies at high risk of bias^1^ (i.e. one or more bias domains were judged high-risk)
- **Inconsistency**: downgrade by one level if heterogeneity was large (I^2^ statistic value >50%, representing potentially substantial heterogeneity)^2^
- **Indirectness**: We will not assess indirectness as patients, interventions and comparators will be similar across studies.
- **Imprecision**: For continuous outcomes, downgrade by one level if the upper or lower limits of the 95% confidence interval crosses the minimal clinically important difference of 10 points ?on a 100-point scale. For dichotomous outcomes. For dichotomous outcomes, downgrade by one level if the lower or upper limits of the 95% confidence interval include appreciable benefit or harm (i.e 95% CI under 0.75 or over 1.25).^4^
- **Publication bias:** downgrade by one level if a funnel plot suggests the presence of publication bias.^5^

## Strategy for data synthesis

All outcome measures will be converted to a 0-100 scale to improve interpretability of findings across studies. We will group findings by condition (spinal pain (low back and neck pain), osteoarthritis (knee, hip), and radicular syndromes (lumbar and cervical)) and antidepressant class and present them as standardised mean differences along with 95% confidence intervals.

If studies are considered to be sufficiently homogenous, results will be pooled. The I^2^ statistics will be used to analyse the between-trial heterogeneity, and a random effects model will be used among trials when I2 > 0%.

## Analysis of subgroups or subsets

If possible, we will perform meta-regression to explore the association of the following factors with treatment effects:

- Risk of bias;
- Small study effects (Less than 100 participants per arm);
- Industry sponsor (yes/no);
- Depression
- Evidence of a dose-response relationship
- Concomitant use of other pain medications

## APPENDIX 1. SEARCH STRATEGIES

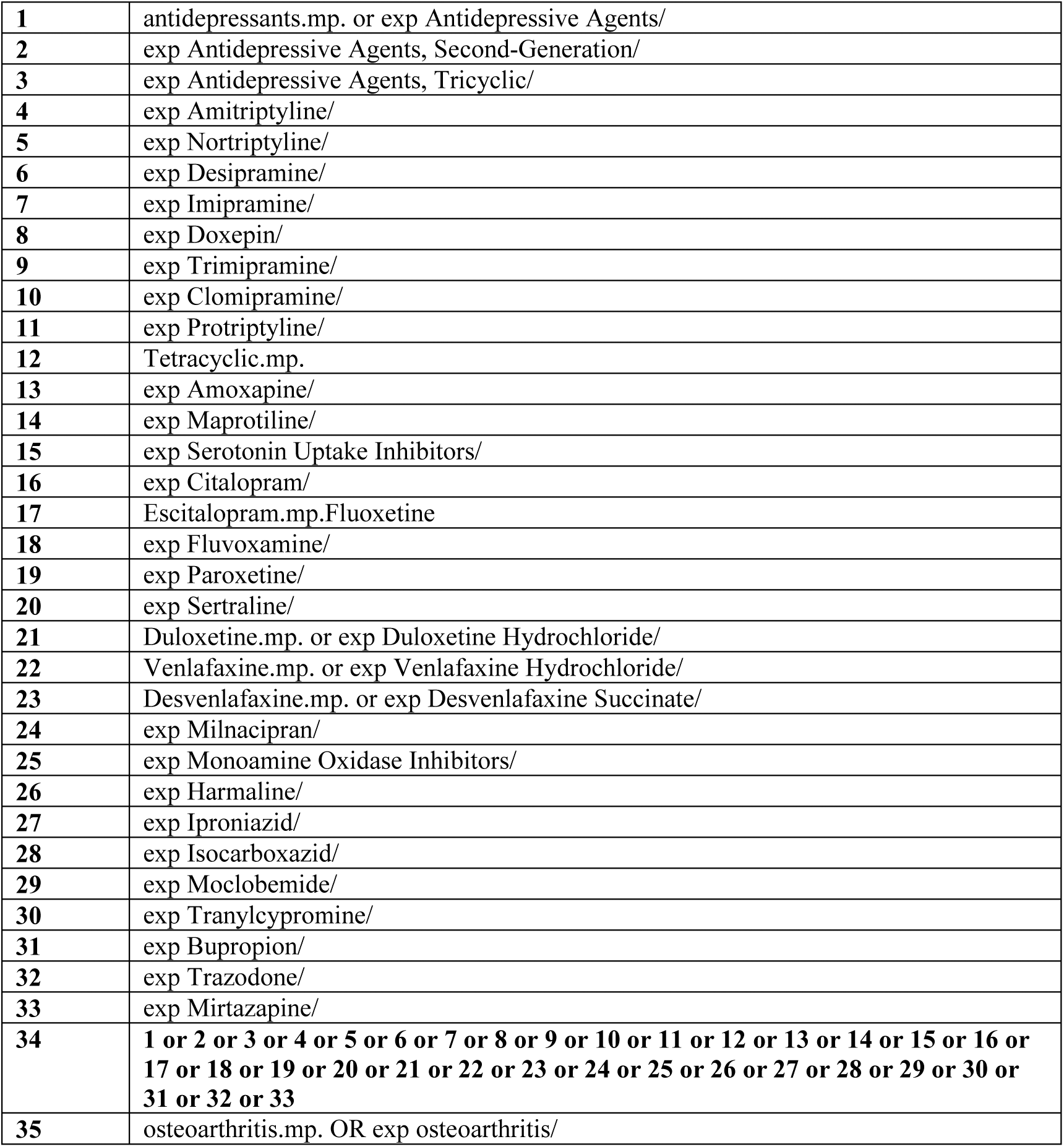
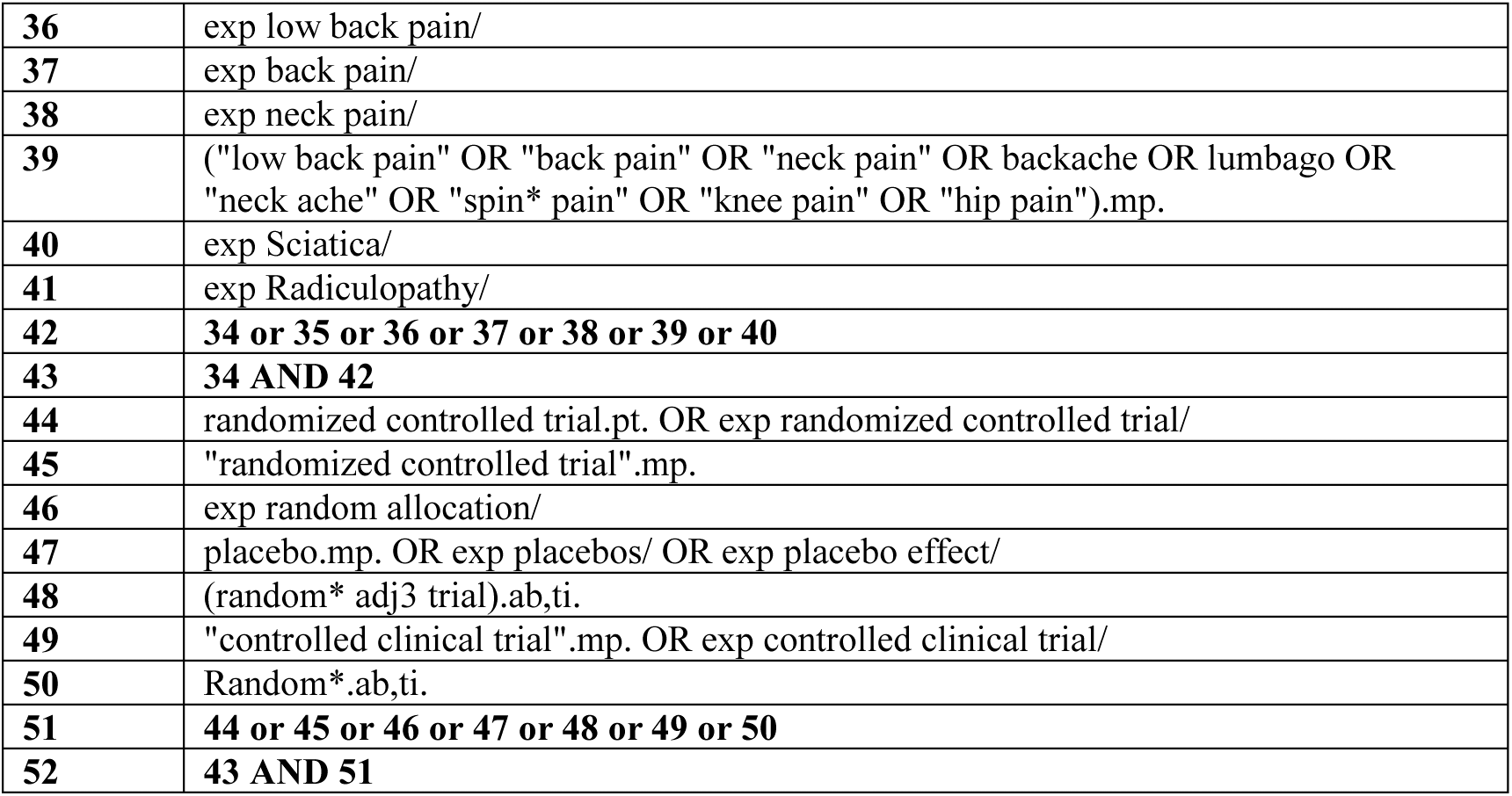

### Pubmed

(((((((Low Back Pain OR back pain OR neck pain OR Sciatica OR Radiculopathy[MeSH Terms])) OR (radiculopathies[Title/Abstract] OR radiculopathy, cervical[Title/Abstract] OR Cervical Radiculopathies[Title/Abstract] OR Cervical Radiculopathy[Title/Abstract] OR Radiculopathies, Cervical[Title/Abstract] OR Nerve Root Disorder[Title/Abstract] OR Radiculitis[Title/Abstract] OR Nerve Root Inflammation[Title/Abstract] OR Compression, Nerve Root[Title/Abstract] OR lumbago[Title/Abstract] OR lower back pain[Title/Abstract] OR low back ache[Title/Abstract] OR low backaches[Title/Abstract] OR postural low back pain[Title/Abstract] OR mechanical low back pain[Title/Abstract] OR recurrent low back pain[Title/Abstract] OR vertebrogenic pain syndrome[Title/Abstract] OR back aches[Title/Abstract] OR back pain with radiation[Title/Abstract] OR neckache[Title/Abstract] OR cervicalgia[Title/Abstract] OR cervicodynias[Title/Abstract] OR cervical pain[Title/Abstract] OR posterior cervical pain[Title/Abstract] OR anterior neck pain[Title/Abstract])) OR (Osteoarthritis OR Osteoarthritis, Spine OR Osteoarthritis, Knee OR Osteoarthritis OR Osteoarthritis, Spine OR Osteoarthritis, Knee[MeSH Terms])) OR (Osteoarthrosis[Title/Abstract] OR Osteoarthroses[Title/Abstract] OR Osteoarthritides[Title/Abstract] OR Arthritis, Degenerative[Title/Abstract] OR Arthritides, Degenerative[Title/Abstract] OR Degenerative Arthritides[Title/Abstract] OR Degenerative Arthritis[Title/Abstract] OR Osteoarthrosis Deformans[Title/Abstract] OR Coxarthrosis[Title/Abstract] OR Coxarthroses[Title/Abstract] OR Osteoarthritis Of Hip[Title/Abstract] OR Osteoarthritis Of Hips[Title/Abstract] OR Knee Osteoarthritides[Title/Abstract] OR Osteoarthritides, Knee[Title/Abstract] OR Osteoarthritis Of Knees [Title/Abstract])))

AND

(((Randomized Controlled Trial [Publication Type] OR Randomized Controlled Trials as Topic OR Controlled Clinical Trial [Publication Type] OR quivalence Trial [Publication Type] OR Random Allocation OR Cross-Over Studies OR Double-Blind Method OR Placebos)) OR (randomization[Title/Abstract] OR Cross-Over Study[Title/Abstract] OR Crossover Studies[Title/Abstract] OR Cross-Over Studies[Title/Abstract] OR Crossover Study[Title/Abstract] OR Crossover Trials[Title/Abstract] OR Crossover design[Title/Abstract] OR Double-Blind Study[Title/Abstract] OR Double-Blind Studies[Title/Abstract] OR allocation[Title/Abstract] OR random[Title/Abstract] OR follow-up[Title/Abstract] OR active placebo[Title/Abstract] OR placebo[Title/Abstract] OR non-inferiority[Title/Abstract] OR superiority[Title/Abstract])))

AND

((((Antidepressive Agents OR Antidepressive Agents, Tricyclic OR Antidepressive Agents, Second-Generation OR Antidepressive Agents, Second-Generation [Pharmacological Action] OR Antidepressive Agents, Tricyclic [Pharmacological Action] OR Adrenergic Uptake Inhibitors OR Antidepressive Agents OR Antidepressive Agents, Tricyclic OR Antidepressive Agents, Second-Generation OR Antidepressive Agents, Second-Generation [Pharmacological Action] OR Antidepressive Agents, Tricyclic [Pharmacological Action] OR Adrenergic Uptake Inhibitors OR))) OR ((Amitriptyline OR Nortriptyline OR Desipramine OR Imipramine OR Doxepin OR Trimipramine OR Clomipramine OR Protriptyline OR Amoxapine OR Maprotiline OR Serotonin Uptake Inhibitors OR Citalopram OR Fluoxetine OR Paroxetine OR Sertraline OR Duloxetine Hydrochloride OR Fluvoxamine OR Venlafaxine Hydrochloride OR Desvenlafaxine Succinate OR Milnacipran OR Monoamine Oxidase Inhibitors OR Harmaline OR Iproniazid OR Isocarboxazid OR Moclobemide OR Tranylcypromine OR Bupropion OR Trazodone OR Mirtazapine[MeSH Terms])))

### EMBASE

|  |  |
| --- | --- |
| 1 | exp knee osteoarthritis/ or exp hip osteoarthritis/ or exp osteoarthritis/ |
| 2 | exp low back pain/ |
| 3 | exp neck pain/ |
| 4 | exp backache/ or exp intervertebral disk hernia/ or exp sciatica/ |
| 5 | exp radiculopathy/ |
| 6 | exp cervicobrachial neuralgia/ |
| 7 | random:.tw. |
| 8 | placebo:.mp. |
| 9 | double-blind:.tw. |
| 10 | antidepressants.mp. or exp antidepressant agent/ |
| 11 | exp amitriptyline/ |
| 12 | exp nortriptyline/ |
| 13 | exp desipramine/ |
| 14 | Exp imipramine/ |
| 15 | exp doxepin/ |
| 16 | exp trimipramine/ |
| 17 | exp clomipramine/ |
| 18 | exp protriptyline/ |
| 19 | exp tetracyclic antidepressant agent/ |
| 20 | exp amoxapine/ |
| 21 | exp maprotiline/ |
| 22 | exp serotonin uptake inhibitor/ |
| 23 | exp citalopram/ |
| 24 | exp fluoxetine/ |
| 25 | exp fluvoxamine/ |
| 26 | exp paroxetine/ |
| 27 | exp sertraline/ |
| 28 | exp duloxetine/ |
| 29 | exp venlafaxine/ |
| 30 | exp desvenlafaxine/ |
| 31 | exp milnacipran/ |
| 32 | exp monoamine oxidase inhibitor/ |
| 33 | exp harmaline/ |
| 34 | exp iproniazid/ |
| 35 | exp isocarboxazid/ |
| 36 | exp moclobemide/ |
| 37 | exp tranylcypromine/ |
| 38 | Bupropion.mp. or exp amfebutamone/ |
| 39 | exp trazodone/ |
| 40 | exp mirtazapine/ |
| 41 | 1 or 2 or 3 or 4 or 5 or 6 |
| 42 | 7 or 8 or 9 |
| 43 | 1 0 or 11 or 12 or 13 or 14 or 15 or 16 or 17 or 18 or 19 or 20 or 21 or 22 or 23 or 24 or 25 or 26 or 27 or 28 or 29 or 30 or 31 or 32 or 33 or 34 or 35 or 36 or 37 or 38 or 39 or 40 |
| 44 | 41 and 42 and 43 |

### Cochrane Central

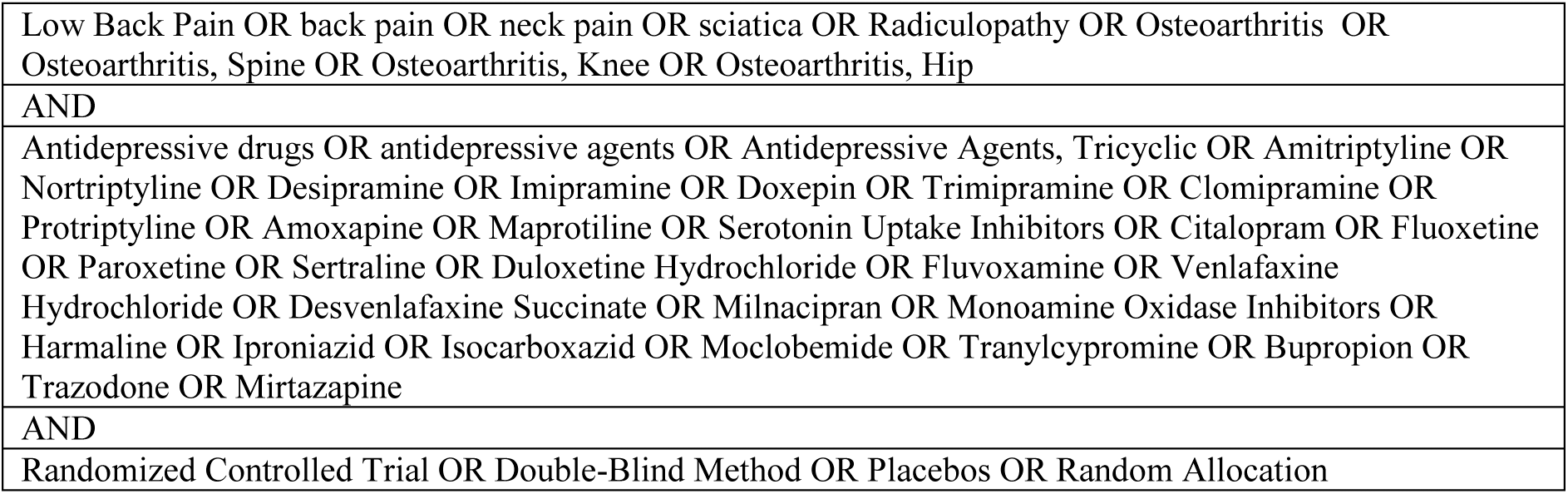

|  |
| --- |
| Low Back Pain OR back pain OR neck pain OR sciatica OR Radiculopathy OR Osteoarthritis OR Osteoarthritis, Spine OR Osteoarthritis, Knee OR Osteoarthritis, Hip |
| AND |
| Antidepressive drugs OR antidepressive agents OR Antidepressive Agents, Tricyclic OR Amitriptyline OR Nortriptyline OR Desipramine OR Imipramine OR Doxepin OR Trimipramine OR Clomipramine OR Protriptyline OR Amoxapine OR Maprotiline OR Serotonin Uptake Inhibitors OR Citalopram OR Fluoxetine OR Paroxetine OR Sertraline OR Duloxetine Hydrochloride OR Fluvoxamine OR Venlafaxine Hydrochloride OR Desvenlafaxine Succinate OR Milnacipran OR Monoamine Oxidase Inhibitors OR Harmaline OR Iproniazid OR Isocarboxazid OR Moclobemide OR Tranlycypromine OR Bupropion OR Trazodone OR Mirtazapine |
| AND |
| Randomized Controlled Trial OR Double-Blind Method OR Placebos OR Random Allocation |

### International Pharmaceutical Abstracts

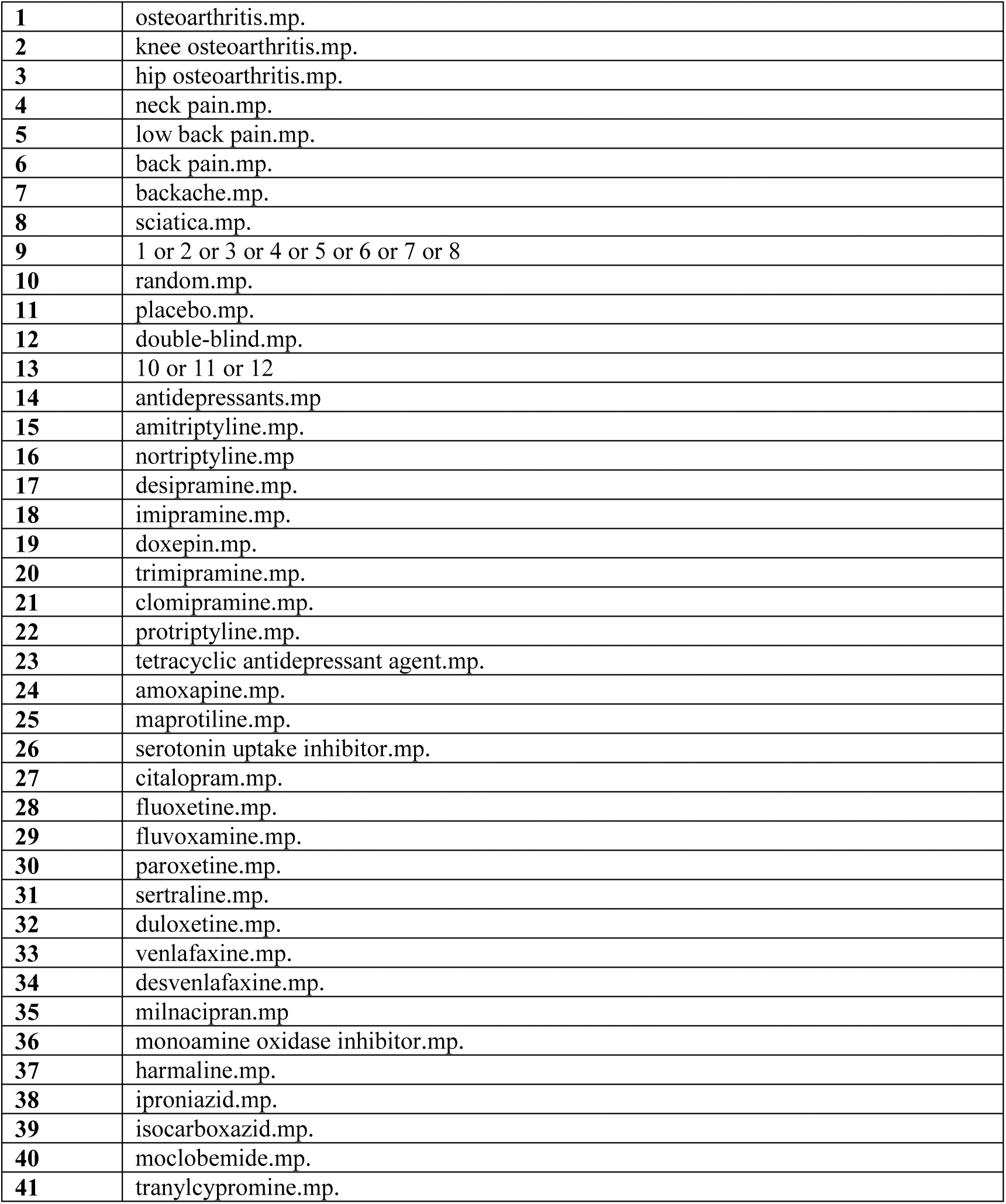
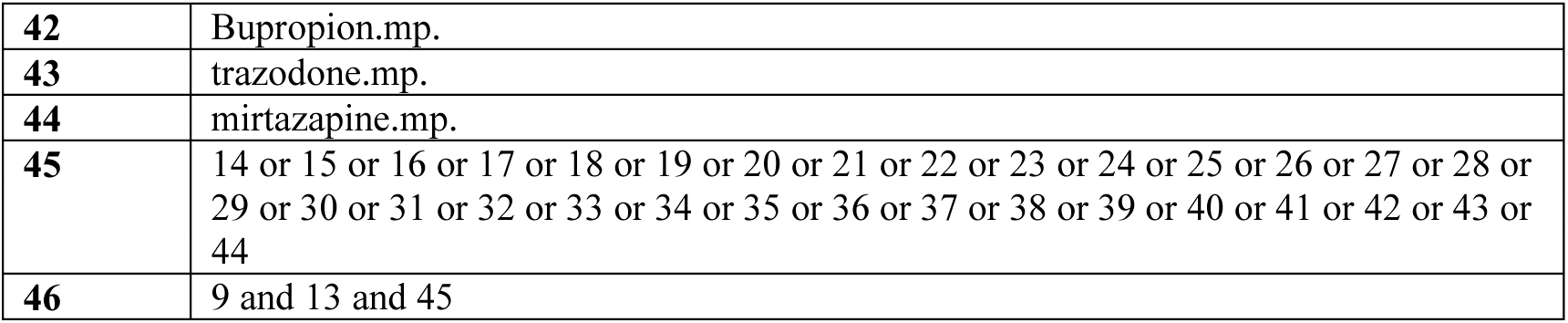

### CINAHL

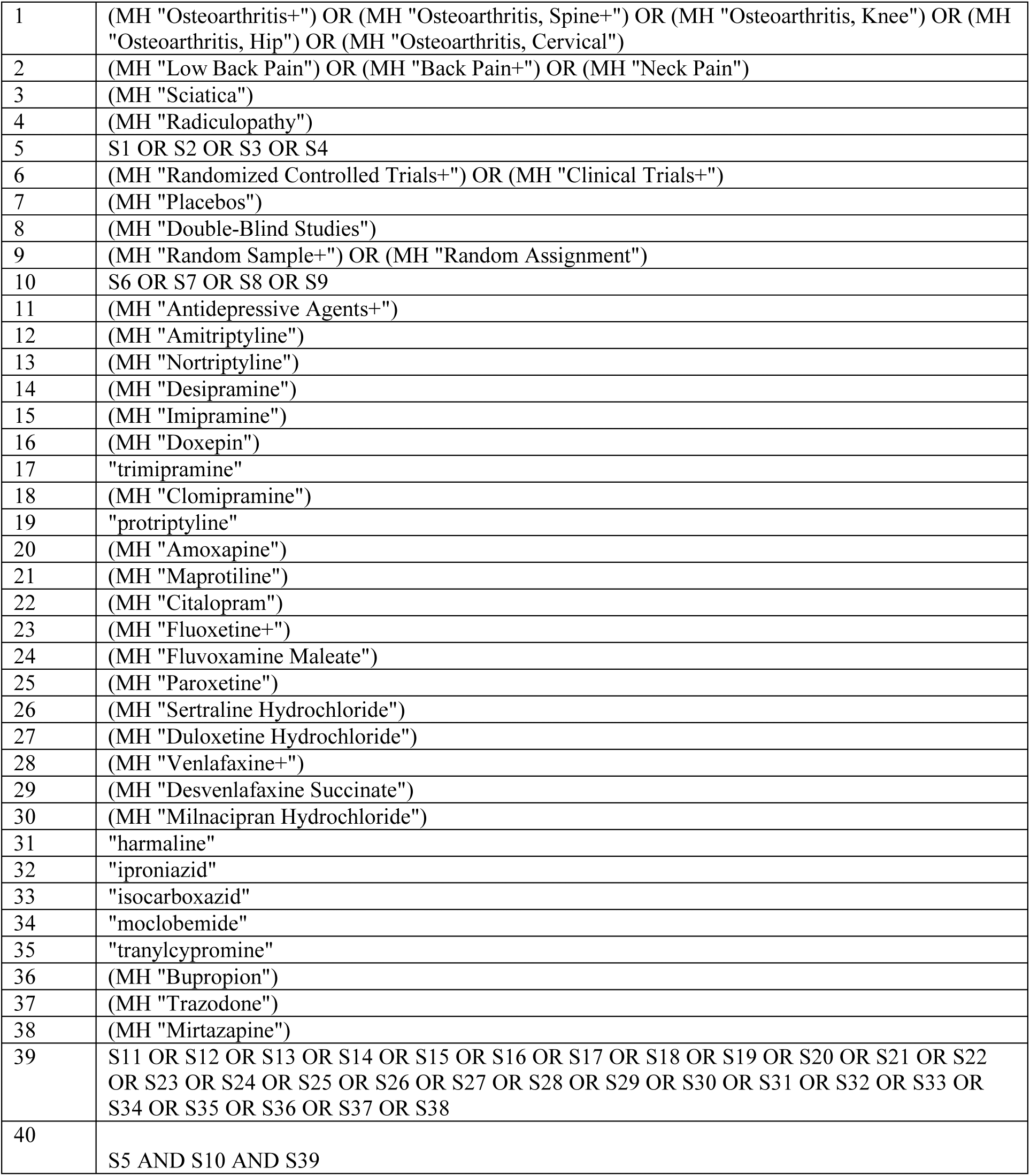

ClinicalTrials.gov and International Clinical Trials Registry Platform*

*Each antidepressant was searched individually

|  |
| --- |
| **1. Low back pain AND** (Amitriptyline OR Nortriptyline OR Desipramine OR Imipramine OR Doxepin OR Trimipramine OR Clomipramine OR protriptyline OR Amoxapine OR Maprotiline OR serotonin uptake inhibitor OR Citalopram OR fluoxetine OR Fluvoxamine OR Paroxetine Sertraline OR Duloxetine OR Venlafaxine OR Desvenlafaxine OR Milnacipran OR Harmaline OR Iproniazid OR Isocarboxazid OR Moclobemide OR Tranylcypromine OR Bupropion OR Trazodone OR mirtazapine) |
| **2. Neck pain AND** (Amitriptyline OR Nortriptyline OR Desipramine OR Imipramine OR Doxepin OR Trimipramine OR Clomipramine OR protriptyline OR Amoxapine OR Maprotiline OR serotonin uptake inhibitor OR Citalopram OR fluoxetine OR Fluvoxamine OR Paroxetine Sertraline OR Duloxetine OR Venlafaxine OR Desvenlafaxine OR Milnacipran OR Harmaline OR Iproniazid OR Isocarboxazid OR Moclobemide OR Tranylcypromine OR Bupropion OR Trazodone OR mirtazapine) |
| **3. Sciatica AND** (Amitriptyline OR Nortriptyline OR Desipramine OR Imipramine OR Doxepin OR Trimipramine OR Clomipramine OR protriptyline OR Amoxapine OR Maprotiline OR serotonin uptake inhibitor OR Citalopram OR fluoxetine OR Fluvoxamine OR Paroxetine Sertraline OR Duloxetine OR Venlafaxine OR Desvenlafaxine OR Milnacipran OR Harmaline OR Iproniazid OR Isocarboxazid OR Moclobemide OR Tranylcypromine OR Bupropion OR Trazodone OR mirtazapine) |
| **4. Osteoarthritis AND** (Amitriptyline OR Nortriptyline OR Desipramine OR Imipramine OR Doxepin OR Trimipramine OR Clomipramine OR protriptyline OR Amoxapine OR Maprotiline OR serotonin uptake inhibitor OR Citalopram OR fluoxetine OR Fluvoxamine OR Paroxetine Sertraline OR Duloxetine OR Venlafaxine OR Desvenlafaxine OR Milnacipran OR Harmaline OR Iproniazid OR Isocarboxazid OR Moclobemide OR Tranylcypromine OR Bupropion OR Trazodone OR mirtazapine) |

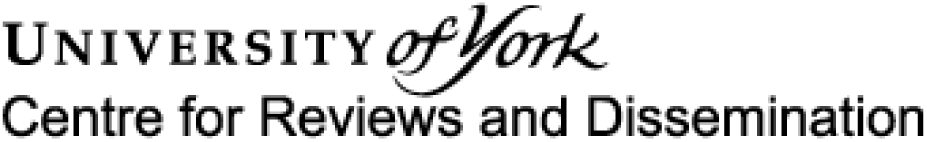

### Systematic review

#### 1. * Review title

Give the working title of the review, for example the one used for obtaining funding. Ideally the title should state succinctly the interventions or exposures being reviewed and the associated health or social problems. Where appropriate, the title should use the PI(E)COS structure to contain information on the Participants, Intervention (or Exposure) and Comparison groups, the Outcomes to be measured and Study designs to be included.

Efficacy and safety of antidepressants for the treatment of spinal pain and osteoarthritis

#### 2. Original language title

For reviews in languages other than English, this field should be used to enter the title in the language of the review. This will be displayed together with the English language title.

Efficacy and safety of antidepressants for the treatment of spinal pain and osteoarthritis

#### 3. * Anticipated or actual start date

Give the date when the systematic review commenced, or is expected to commence.

01/12/2019

#### 4. * Anticipated completion date

Give the date by which the review is expected to be completed.

12/05/2020

#### 5. * Stage of review at time of this submission

Indicate the stage of progress of the review by ticking the relevant Started and Completed boxes. Additional information may be added in the free text box provided.

Please note: Reviews that have progressed beyond the point of completing data extraction at the time of initial registration are not eligible for inclusion in PROSPERO. Should evidence of incorrect status and/or completion date being supplied at the time of submission come to light, the content of the PROSPERO record will be removed leaving only the title and named contact details and a statement that inaccuracies in the stage of the review date had been identified.

This field should be updated when any amendments are made to a published record and on completion and publication of the review. If this field was pre-populated from the initial screening questions then you are not able to edit it until the record is published.

The review has not yet started: Yes

| Review stage | Started | Completed |
| --- | --- | --- |
| Preliminary searches | No | No |
| Piloting of the study selection process | No | No |
| Formal screening of search results against eligibility criteria | No | No |
| Data extraction | No | No |
| Risk of bias (quality) assessment | No | No |
| Data analysis | No | No |

Provide any other relevant information about the stage of the review here (e.g. Funded proposal, protocol not yet finalised).

#### 6. * Named contact

The named contact acts as the guarantor for the accuracy of the information presented in the register record.

Giovanni Ferreira

### Email salutation (e.g. “Dr Smith” or “Joanne”) for correspondence

Mr Ferreira

#### 7. * Named contact email

Give the electronic mail address of the named contact.

#### 8. Named contact address

Give the full postal address for the named contact.

Level 10 North KGV V Building Missenden Road, Camperdown, NSW, 2050, Sydney, Australia

#### 9. Named contact phone number

Give the telephone number for the named contact, including international dialling code.

+6186276691

#### 10. * Organisational affiliation of the review

Full title of the organisational affiliations for this review and website address if available. This field may be completed as ‘None’ if the review is not affiliated to any organisation.

The University of Sydney

### Organisation web address

#### 11. * Review team members and their organisational affiliations

Give the personal details and the organisational affiliations of each member of the review team. Affiliationrefers to groups or organisations to which review team members belong. NOTE: email and country are now mandatory fields for each person.

Mr Giovanni Ferreira. The University of Sydney

Assistant/Associate Professor Christine Lin. The University of Sydney

Dr Joshua Zadro. The University of Sydney

Dr Christina Abdel-Shaheed. The University of Sydney

Professor Andrew McLachlan. The University of Sydney

Dr Mary O’Keeffe. The University of Sydney

Professor Chris Maher. The University of Sydney

#### 12. * Funding sources/sponsors

Give details of the individuals, organizations, groups or other legal entities who take responsibility for initiating, managing, sponsoring and/or financing the review. Include any unique identification numbers assigned to the review by the individuals or bodies listed.

None

### Grant number(s)

#### 13. * Conflicts of interest

List any conditions that could lead to actual or perceived undue influence on judgements concerning the main topic investigated in the review.

None

#### 14. Collaborators

Give the name and affiliation of any individuals or organisations who are working on the review but who are not listed as review team members. **NOTE: email and country are now mandatory fields for each person**.

#### 15. * Review question

State the question(s) to be addressed by the review, clearly and precisely. Review questions may be specific or broad. It may be appropriate to break very broad questions down into a series of related more specific questions. Questions may be framed or refined using PI(E)COS where relevant.

**Are antidepressants efficacious and safe for patients with spinal pain or osteoarthritis?**

#### 16. * Searches

State the sources that will be searched. Give the search dates, and any restrictions (e.g. language or publication period). Do NOT enter the full search strategy (it may be provided as a link or attachment.)

#### 17. URL to search strategy

Give a link to a published pdf/word document detailing either the search strategy or an example of a search strategy for a specific database if available (including the keywords that will be used in the search strategies), or upload your search strategy. Do NOT provide links to your search results.

Alternatively, upload your search strategy to CRD in pdf format. Please note that by doing so you are consenting to the file being made publicly accessible.

Do not make this file publicly available until the review is complete

#### 18. * Condition or domain being studied

Give a short description of the disease, condition or healthcare domain being studied. This could include health and wellbeing outcomes.

Spinal pain (low back pain and/or neck pain), and hip and/or knee osteoarthritis (OA).

#### 19. * Participants/population

Give summary criteria for the participants or populations being studied by the review. The preferred format includes details of both inclusion and exclusion criteria.

#### 20. * Intervention(s), exposure(s)

Give full and clear descriptions or definitions of the nature of the interventions or the exposures to be reviewed.

Antidepressants are recommended by clinical practice guidelines for low back pain and hip and knee osteoarthritis. We will include trials testing any type of antidepressant drug prescribed at any dose, as treatment for spinal pain and/or hip and/or knee OA. Studies testing drug combinations (e.g. an antidepressant in addition to another analgesic) will only be included if the comparison treatment is the same analgesic and placebo. Types of antidepressants drugs to be included in this review include, but are not limited to:

Tricyclic antidepressants (e.g. amitriptyline, nortriptyline, desipramine, imipramine, doxepin, dothiepin, trimipramine, clomipramine and protriptyline)

- Tetracyclic antidepressants (e.g. mianserin amoxapine and maprotiline)
- Selective serotonin reuptake inhibitors (SSRIs) (e.g. citalopram, escitalopram, fluoxetine, fluvoxamine, paroxetine, Zimelidine and sertraline)
- Serotonin and norepinephrine reuptake inhibitors (SNRIs) (e.g. Duloxetine, Venlafaxine, Desvenlafaxine, Milnacipran)
- Monoamine oxidase inhibitors (MAOIs) (e.g. harmaline, iproclozide, iproniazid, isocarboxazid, moclobemide nialamide, toloxatone and tranylcypromine)
- Atypical antidepressants (e.g. Bupropion, Trazodone, Mirtazapine, Nefazodone, agomelatine)

#### 21. * Comparator(s)/control

Where relevant, give details of the alternatives against which the main subject/topic of the review will be compared (e.g. another intervention or a non-exposed control group). The preferred format includes details of both inclusion and exclusion criteria.

#### 22. * Types of study to be included

Give details of the types of study (study designs) eligible for inclusion in the review. If there are no restrictions on the types of study design eligible for inclusion, or certain study types are excluded, this should be stated. The preferred format includes details of both inclusion and exclusion criteria.

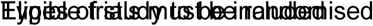 placebo-controlled trials published as full-text in peer-reviewed journals. Both enriched and non-enriched designs will be included. We will contact authors who have published a protocol for a relevant trial that has not yet been published in an attempt to access the data or to obtain a copy of the publication. There will be no restrictions for language or publication date. We will attempt to translate studies published in languages other than English.

#### 23. Context

Give summary details of the setting and other relevant characteristics which help define the inclusion or exclusion criteria.

#### 24. * Main outcome(s)

Give the pre-specified main (most important) outcomes of the review, including details of how the outcome is defined and measured and when these measurement are made, if these are part of the review inclusion criteria.

2. 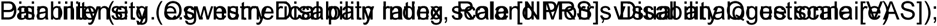

3. Composite disease scores (e.g Western Ontario and McMaster Universities osteoarthritis index [WOMAC], Knee Osteoarthritis Outcome Score [KOOS], Knee Society Scoring System [KSSS], Lysholm Knee Scoring Scale [LKSC], Hip Osteoarthritis Outcome Score [HOOS], Harris Hip Score [HHS])

### * Measures of effect

Please specify the effect measure(s) for you main outcome(s) e.g. relative risks, odds ratios, risk difference, and/or ‘number needed to treat.

2. 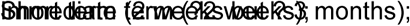

3. Intermediate term (3 months but ?12 months);

4. Long term (12 months).

#### 25. * Additional outcome(s)

List the pre-specified additional outcomes of the review, with a similar level of detail to that required for main outcomes. Where there are no additional outcomes please state ‘None’ or ‘Not applicable’ as appropriate to the review

1 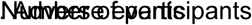 reporting any adverse events (as defined by each study)

• Number of participants reporting any serious adverse events (as defined by each study)

2. Number (%) of participants who stopped the study medicine due to adverse effects

3. Number (%) of participants who stopped the study medicine due to lack of efficacy

4. Number (%) of participants who withdrew from study citing either of above

5. Number (%) of participants who were lost to follow-up

### • Measures of effect

Please specify the effect measure(s) for you additional outcome(s) e.g. relative risks, odds ratios, risk difference, and/or ‘number needed to treat.

2. 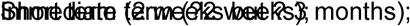

3. Intermediate term (3 months but ?12 months);

4. Long term (12 months).

#### 26. * Data extraction (selection and coding)

Describe how studies will be selected for inclusion. State what data will be extracted or obtained. State how this will be done and recorded.

#### 27. * Risk of bias (quality) assessment

Describe the method of assessing risk of bias or quality assessment. State which characteristics of the studies will be assessed and any formal risk of bias tools that will be used.

We will assess risk of bias using the Cochrane RoB tool and assess the quality of evidence with GRADE. For each GRADE domain, the quality of evidence will be downgraded by one level if a serious flaw was present 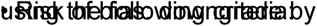 one level if more than 25% of participants are from studies at high risk of bias (i.e. one or more bias domains were judged high-risk)

- Inconsistency: downgrade by one level if heterogeneity was large (I2 statistic value 50%, representing potentially substantial heterogeneity)
- Indirectness: We will not assess indirectness as patients, interventions and comparators will be similar across studies.
- Imprecision: For continuous outcomes, downgrade by one level if the limits of the 95% confidence interval are 20 points different to the point estimate; i.e. twice the minimal clinically important difference of 10 points on a 100-point scale. For dichotomous outcomes, downgrade by one level if the lower or upper limits of the 95% confidence interval include appreciable benefit or harm (i.e. 95% CI under 0.75 or over 1.25).
- Publication bias: downgrade by one level if a funnel plot suggests the presence of publication bias.

#### 28. * Strategy for data synthesis

Provide details of the planned synthesis including a rationale for the methods selected. This **must not be generic text** but should be **specific to your review** and describe how the proposed analysis will be applied to your data.

All outcome measures will be converted to a 0-100 scale to improve interpretability of findings across studies. We will group findings by condition (spinal pain (low back and neck pain), osteoarthritis (knee, hip), and radicular syndromes (lumbar and cervical) and antidepressant class and present them as standardised 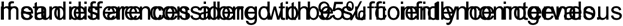, results will be pooled. The I^2^ statistics will be used to analyse the between-trial heterogeneity, and a random effects model will be used among trials when I^2^ 0%.

#### 29. * Analysis of subgroups or subsets

State any planned investigation of ‘subgroups’. Be clear and specific about which type of study or participant will be included in each group or covariate investigated. State the planned analytic approach.

If possible, we will perform meta-regression to explore the association of the following factors with treatment 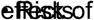 bias;

- Small study effects (Less than 100 participants per arm);
- Industry sponsor (yes/no);
- Depression
- Evidence of a dose-response relationship
- Concomitant use of other pain medications

#### 30. * Type and method of review

Select the type of review and the review method from the lists below. Select the health area(s) of interest for your review.

Type of review
Cost effectiveness No Diagnostic No Epidemiologic No Individual patient data (IPD) meta-analysis No Intervention Yes Meta-analysis Yes Methodology No Narrative synthesis No Network meta-analysis No Pre-clinical No Prevention No Prognostic No Prospective meta-analysis (PMA) No Review of reviews No Service delivery No Synthesis of qualitative studies No Systematic review Yes Other No

Health area of the review
Alcohol/substance misuse/abuse No Blood and immune system No Cancer No Cardiovascular No Care of the elderly No Child health No Complementary therapies No Crime and justice No Dental No Digestive system No Ear, nose and throat No Education No Endocrine and metabolic disorders No Eye disorders No General interest No Genetics No Health inequalities/health equity No Infections and infestations No International development No Mental health and behavioural conditions No Musculoskeletal Yes Neurological No Nursing No Obstetrics and gynaecology No Oral health No Palliative care No Perioperative care No Physiotherapy No Pregnancy and childbirth No Public health (including social determinants of health) No Rehabilitation No Respiratory disorders No Service delivery No Skin disorders No Social care No Surgery No Tropical Medicine No Urological No Wounds, injuries and accidents No Violence and abuse No

#### 31. Language

Select each language individually to add it to the list below, use the bin icon to remove any added in error. English

There is not an English language summary

#### 32. * Country

Select the country in which the review is being carried out from the drop down list. For multi-national collaborations select all the countries involved.

Australia

#### 33. Other registration details

Give the name of any organisation where the systematic review title or protocol is registered (such as with The Campbell Collaboration, or The Joanna Briggs Institute) together with any unique identification number assigned. (N.B. Registration details for Cochrane protocols will be automatically entered). If extracted data will be stored and made available through a repository such as the Systematic Review Data Repository (SRDR), details and a link should be included here. If none, leave blank.

#### 34. Reference and/or URL for published protocol

Give the citation and link for the published protocol, if there is one

Give the link to the published protocol.

Alternatively, upload your published protocol to CRD in pdf format. Please note that by doing so you are consenting to the file being made publicly accessible.

Yes I give permission for this file to be made publicly available

Please note that the information required in the PROSPERO registration form must be completed in full even if access to a protocol is given.

#### 35. Dissemination plans

Give brief details of plans for communicating essential messages from the review to the appropriate audiences.

Do you intend to publish the review on completion?

Yes

#### 36. Keywords

Give words or phrases that best describe the review. Separate keywords with a semicolon or new line. Keywords will help users find the review in the Register (the words do not appear in the public record but are included in searches). Be as specific and precise as possible. Avoid acronyms and abbreviations unless these are in wide use.

Low back pain, neck pain, sciatica, antidepressants, pain

#### 37. Details of any existing review of the same topic by the same authors

Give details of earlier versions of the systematic review if an update of an existing review is being registered, including full bibliographic reference if possible.

#### 38. * Current review status

Review status should be updated when the review is completed and when it is published. For newregistrations the review must be Ongoing.

Please provide anticipated publication date

Review_Ongoing

#### 39. Any additional information

Provide any other information the review team feel is relevant to the registration of the review.

#### 40. Details of final report/publication(s)

This field should be left empty until details of the completed review are available.

Give the link to the published review.

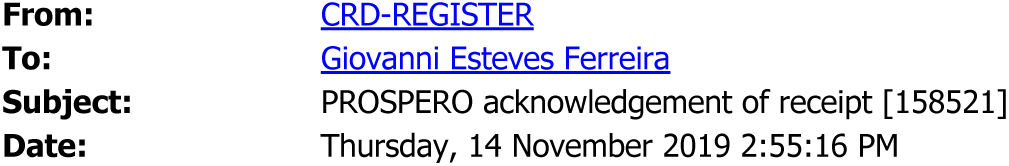

Dear Registrant,

Thank you for submitting details of your systematic review for registration in PROSPERO.

We will check the information supplied to

- make sure that your systematic review is within scope
- ensure that the fields have been completed appropriately.

PLEASE NOTE THAT THESE CHECKS DO NOT CONSTITUTE PEER REVIEW OR IMPLY APPROVAL OF YOUR SYSTEMATIC REVIEW METHODS.

We will let you know when your record has been published on PROSPERO, or alternatively ask for further information or clarification. If your application is rejected we will advise you of the reasons for non-publication (usually this will be if your review is out of scope).

With the current extremely high demand for registration, we will aim to respond within 10 working days for UK submissions but for submissions from outside the UK it will be considerably longer - possibly around three months.

But we will process your application as soon as possible. During this time the record will be locked and you will not be able to access it.

Please note that this does not stop you working on your review.

Yours sincerely,

PROSPERO Administrator

Centre for Reviews and Dissemination

University of York

York YO10 5DD

t: +44 (0) 1904 321049

e

www.york.ac.uk/inst/crd

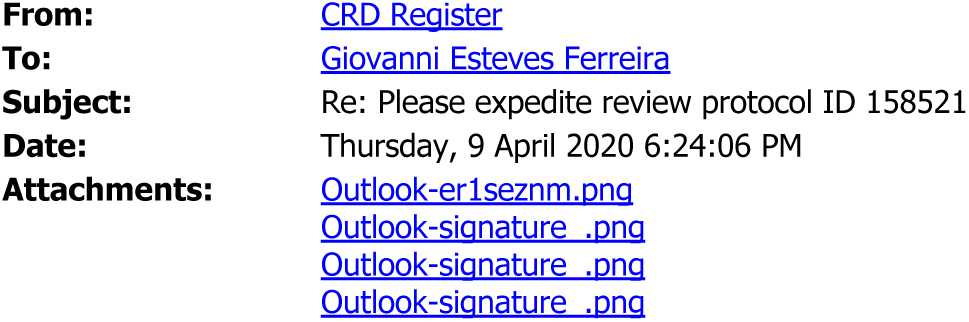

We have received your submission.

But we have experienced an unprecedented number of submissions for registration this past year and consequently there is a substantial delay in processing records at the moment. We are currently giving priority to COVID-19 studies, and still aim to respond within 10 <u>working</u> days for UK submissions, but for submissions from the rest of the world it will be considerably longer (now over five months).

We are a small team (the equivalent of 2.5 full time staff) processing submissions from all over the world and demand is currently EXTREMELY high - we have over 13000 records in the queue for processing! Remember that it is a service that is free to you.

We will check the information supplied to

- make sure that your systematic review is within scope
- ensure that the fields have been completed appropriately.

Please note that these checks

**Do not constitute peer review**

or imply **approval of your systematic review methods**.

Waiting to hear from us does not prevent you from progressing with your review.

Other sites where you can register your review include Research Registry https://www.researchregistrv.com/

Open Science Framework https://osf.io/

**PLEASE NOTE: Failure to comply with our registration requirements will result in delays in your registration process. Our workload is ever increasing and if we send your record back for amendment, it then goes to the end of the queue. PLEASE READ OUR GUIDANCE CAREFULLY and complete the form correctly to prevent further delay**.

See https://www.crd.york.ac.uk/PROSPERO/#aboutregpage

PROSPERO Admin team

Centre for Reviews and Dissemination

University of York

York

YO10 5DD

t: +44 (0) 1904 321049

www.york.ac.uk/inst/crd

www.crd.york.ac.uk/prospero/

CRD is a department of the University of York.

EMAIL DISCLAIMER: http://www.york.ac.uk/docs/disclaimer/email.htm

On Thu, 9 Apr 2020 at 06:33, Giovanni Esteves Ferreira wrote:

To whom it may concern,

this email is in regards to the systematic review “Efficacy and safety of antidepressants for the treatment of spinal pain and osteoarthritis” registered on PROSPERO on November 14th 2019. I have registered the review almost five months ago and have not yet received a confirmation of the register. As I am close to submitting the systematic review I wonder if wouldn’t it be possible to expedite the registration.

I understand you staff is usually overwhelmed by the great number of submissions, and I can imagine you have been receiving countless review protocols focusing on COVID-19. But it would be much appreciated if my review could be assessed soon.

I look forward to hearing from you.

Giovanni

Giovanni Ferreira | PhD candidate The University of Sydney

Faculty of Medicine and Health, Sydney School of Public Health, Institute for Musculoskeletal Health Level 10 KGV Building | Missenden Road, Camperdown | NSW | 2050 T +61 2 8627 6691 | M +61 410 531 601 | @qiovanni_ef

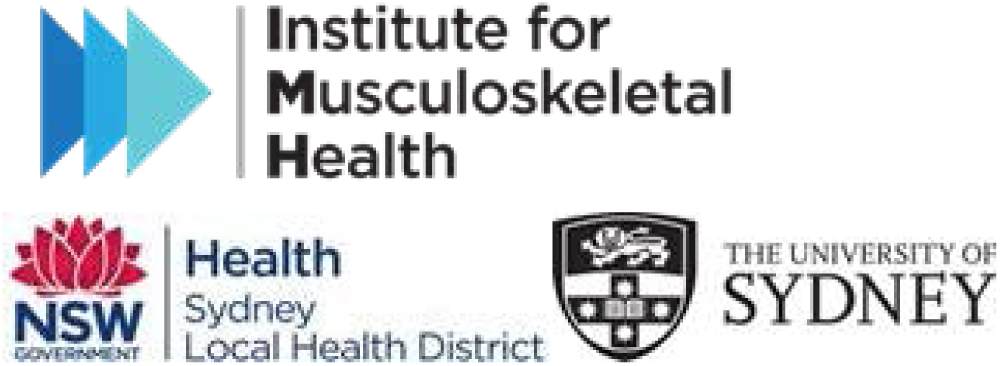

## References

1. Oliveira CB, Maher CG, Pinto RZ, et al. Clinical practice guidelines for the management of non-specific low back pain in primary care: an updated overview. Eur Spine J. 2018;27(11):2791–2803.

2. Bannuru RR, Osani MC, Vaysbrot EE, et al. OARSI guidelines for the non-surgical management of knee, hip, and polyarticular osteoarthritis. Osteoarthritis Cartilage. 2019;27(11):1578–1589.

3. Royal Australian College of General Practitioners. Guideline for the management of knee and hip osteoarthritis. 2nd ed. 2018.

## References

1. Guyatt GH, Oxman AD, Vist G, et al. GRADE guidelines: 4. Rating the quality of evidence--study limitations (risk of bias). J Clin Epidemiol. 2011;64(4):407–415.

2. Guyatt GH, Oxman AD, Kunz R, et al. GRADE guidelines: 7. Rating the quality of evidence--inconsistency. J Clin Epidemiol. 2011;64(12):1294–1302.

3. Guyatt GH, Oxman AD, Kunz R, et al. GRADE guidelines: 8. Rating the quality of evidence--indirectness. J Clin Epidemiol. 2011;64(12):1303–1310.

4. Guyatt GH, Oxman AD, Kunz R, et al. GRADE guidelines 6. Rating the quality of evidence-- imprecision. J Clin Epidemiol. 2011;64(12):1283–1293.

5. Guyatt GH, Oxman AD, Montori V, et al. GRADE guidelines: 5. Rating the quality of evidence-publication bias. J Clin Epidemiol. 2011;64(12):1277–1282

